# Urinary Astrocyte-derived Extracellular Vesicles: A Non-invasive Tool for Capturing Human *In Vivo* Molecular “Movies” of Brain

**DOI:** 10.1101/2024.01.12.24301104

**Authors:** Xin-hui Xie, Mian-mian Chen, Shu-xian Xu, Jun-hua Mei, Qing Yang, Chao Wang, Zhongchun Liu

## Abstract

The identification of particularly individual-level biomarkers, for certain central nervous system (CNS) diseases remains challenging. A recent approach involving the enrichment of brain-derived extracellular vesicles (BDEVs) from peripheral blood has emerged as a promising method to obtain direct *in vivo* CNS data, bypassing the blood-brain barrier. However, for rapidly evolving CNS diseases (*e.g.*, weeks or even days), the Nyquist-Shannon sampling theorem dictates the need for a high-frequency sampling rate. Obviously, daily collection of blood or cerebrospinal fluid from human subjects is impractical. Thus, we innovated a novel method to isolate astrocyte-derived EVs from urine (uADEVs). It involves three main steps: 1) concentrating urine samples, 2) isolating total EVs from urine (uTEVs) using ultracentrifugation, and 3) using an anti-glutamate/aspartate transporter (GLAST) antibody to isolate GLAST^+^EVs from uTEVs. Subsequently, we confirmed the identity of these GLAST^+^EVs as uADEVs using transmission electron microscopy, nanoparticle tracking analysis, western blotting, and the measurement of astrocyte-related neurotrophins. Furthermore, we applied the uADEVs protocol to depict the detailed trajectory of the N-methyl-d-aspartic acid receptor (NMDAR) subunit zeta-1 (GluN1) in an anti-NMDAR encephalitis patient, demonstrated the potential of this method for capturing intricate trajectories of CNS-specific molecular *in vivo* signals at the individual level. This non-invasive approach enables frequent sampling, up to daily or even half-daily, analogous to capturing molecular “movies” of the brain, coupled with appropriate signal processing algorithms, holds promise for identifying novel biomarkers or illuminating the etiology of rapidly evolving CNS diseases by tracking the precise trajectories of target molecules.

## Introduction

Research on central nervous system (CNS) diseases, especially mental disorders like schizophrenia, depression, and bipolar disorder, has been gradual, and reliable biological markers remain unidentified^1,2^. Several key factors may contribute to this dilemma.

To begin, the CNS boasts distinctive attributes, notably the presence of the blood-brain barrier (BBB), which poses a direct hurdle to identifying CNS abnormalities^3^. This challenge is compounded by the BBB’s high selectivity, leading to marked inconsistencies in the expression levels of molecules within the CNS compared to the periphery^4–7^. Such incongruities substantially complicate research endeavors. Additionally, the dearth of expedient and non-invasive sampling techniques is notable. The most direct sampling method for the CNS is brain biopsy, but this is clearly very difficult to perform. Another option, cerebrospinal fluid (CSF) collection via lumbar puncture, is neither unsuitable for routine application. While peripheral blood is the most commonly employed biological sample, its representation of the CNS is limited^4,8,9^.

Furthermore, a paucity in sampling frequency is evident. Although reliable biomarkers have been identified for specific neurological and psychiatric disorders like Alzheimer’s disease (AD) and Parkinson’s disease (PD)—Aβ^10,11^, tau^12,13^, and α-synuclein^14^. These biomarkers greatly expedited research in these domains, but a tacit assumption should not be ignored: these disorders exhibit protracted disease courses. Adopting a wave-based perspective, these biomarkers’ trajectories resemble long waves, with wavelengths spanning years or decades. Therefore, according to the Nyquist-Shannon sampling theorem^15^, detecting these extended biomarker waves necessitates lower frequencies, rendering half-yearly or yearly sampling adequate. However, for disorders characterized by rapid fluctuations—such as depression, bipolar disorder, encephalitis, and others—exhibiting weekly, daily, or even hourly changes, routine follow-up intervals (monthly, half-yearly, or yearly) fall short of capturing the molecular trajectories which aligned with symptom progression. And since daily sampling of peripheral blood or CSF is impractical, there is a need for a new methodological approach that can quickly and non-invasively explore the CNS and facilitate research on these rapidly changing neurological and psychiatric diseases. We focused on extracellular vesicles (EVs) as a potential tool for this purpose.

EVs are found in various bodily fluids, including blood, urine, tears, and saliva^16^, and have emerged as promising tools for identifying disease biomarkers, serving as liquid biopsies^17^. Notably, EVs can cross the BBB bidirectionally^18^, making brain-derived EVs (BDEVs) a potential “window to the brain”^19^. In a large-sample trial, the concentrations of T-tau, P-T181-Tau, and Aβ_1-42_ in serum neuro-derived EVs (NDEVs) were linearly associated with their concentrations in CSF with correlation coefficients close to 0.9^20^. Furthermore, these NDEVs were able to predict the onset of AD^21^. Similarly, in the case of PD, α-synuclein, a biomarker of PD, was found to be elevated in NDEVs in PD patients^22–26^, and the area under the receiver operating characteristic curve (ROC) exceeded 0.9^27^. Additionally, animal studies also shown a high level of consistency between plasma astrocyte-derived EVs (ADEVs) and brain homogenous (BH)^28^. In short, the plasma/serum BDEVs could be good proxies of CNS^29,30^. However, the collection of peripheral blood is also an invasive procedure that is impractical for daily sampling, limiting the sampling rate for obtaining *in vivo* signals from the CNS using plasma/serum BDEVs. Additionally, the presence of heteroproteins in peripheral blood makes it challenging and inconvenient to isolate EVs from specific cell sources.

To bypass the disadvantages of isolating BDEVs from peripheral blood, we focused on another type of body fluid—urine, which also contains a large amount of EVs^31^. Urine is an optimal body fluid for identifying diagnostic biomarkers due to its capacity for large-scale and high-frequency collection, as well as its non-invasive nature. Urinary EVs (uEVs) have been implicated in the pathophysiological mechanisms of urogenital diseases and hold potential as molecular biomarkers for these conditions^32–35^. Initially, uEVs were thought to originate primarily from cells in the urogenital tract, including the kidneys, bladder, and sex glands^36^. However, given that primary urine results from plasma filtration in the glomeruli, EVs in blood might enter and be detected in urine^37,38^. For example, the labeled EVs were injected intravenously into rats, and later found in their urine^39^. Wang et al. identify neuronal marker protein in urinary total EVs (uTEVs)^35^, and Fraser et al. reported elevated levels of ser(P)-1292 LRRK2, a PD-associated protein, in uTEVs of PD patients, correlating with cognitive and daily function impairments^32^. It suggests a possibility that non-urogenital EVs, including BDEVs, can enter urine and be isolated. As EVs are considered to reflect the state of their origin cells, and urine is a readily accessible and non-invasive biofluid, the successful isolation of specific EVs from urine could therefore serve as a valuable tool for diagnostic and physiological research.

Thus, here we developed a protocol that enables the enrichment of the glutamate/aspartate transporter (GLAST)^+^EVs which is believed to be ADEVs^40–47^ from urine, namely urinary ADEVs (uADEVs). We believe that uADEVs could serve as a valuable tool for non-invasive, high-frequency daily sampling of human *in vivo* CNS signals, enabling the collection of large-scale longitudinal data on the dynamic behavior of these cells.

## 2. Materials and Methods

This study was conducted at Renmin Hospital of Wuhan University (Mental Health Center of Hubei Province, Wuhan, Hubei, China) and Wuhan First Hospital in compliance with the Declaration of Helsinki (revised edition, 2013)^48^. The study protocol was approved by both the Human Ethics Committee of Renmin Hospital of Wuhan University and Wuhan First Hospital. All participants provided informed consent and were free to withdraw from the trial at any time for any reason.

### 2.1 Isolation protocol of uADEVs

Generally, in this protocol, we first concentrated the urine samples and isolated the uTEVs using ultracentrifugation (UC), followed by the isolation of uADEVs using biotin-anti-GLAST-antibody, similar to the isolation of ADEVs from plasma or serum^40–47,49^. The flow chart of this protocol is depicted in **Figure 1(a)**.

**Figure 1.**
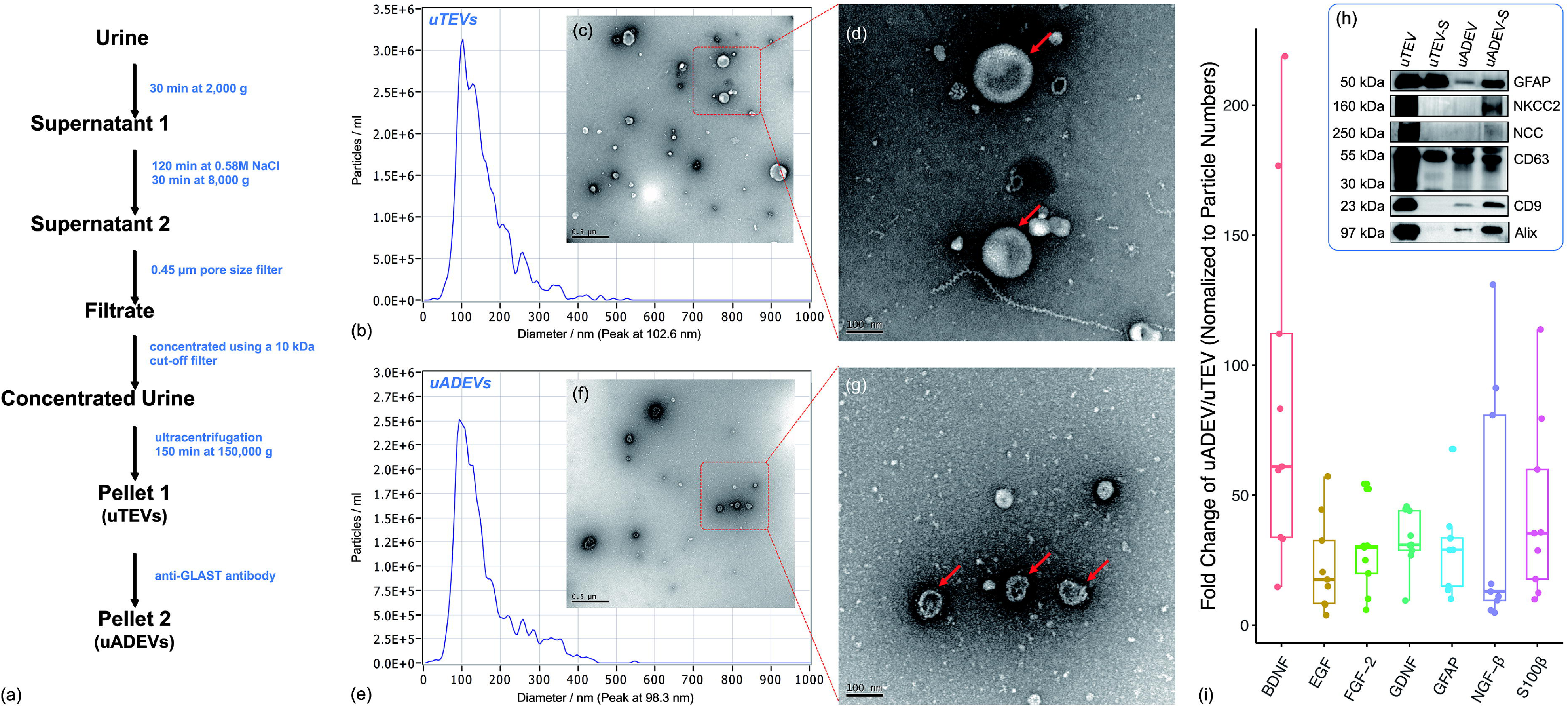
Isolation and Validations of uADEVs. (a) Schematic diagram of the uADEVs isolation protocol. (b and e) NTA results of uTEVs and uADEVs. (c, d, f, and g) TEM images of uTEVs and uADEVs (scale bars: 0.5 μm and 100nm). (h) Results of western blotting: Three EV markers (CD63, CD9 and Alix) and an astrocyte marker (GFAP) were present in the ADEVs sample, while two kidney markers (NKCC2 and NCC) were absent. (i) Significantly increased astrocyte-related neurotrophic factors in uADEVs.)

#### 2.1.1 Isolation of uTEV

Nine healthy volunteers, comprising six males and three females, participated in the study. The median age of the participants was 25.0 years with an interquartile range (IQR) of 4.0 years. A total of 300–600 ml of fresh morning urine of each participant was collected and promptly delivered to the laboratory. The samples were processed within two hours of collection. The urine sample was centrifuged at room temperature (RT) for 30 minutes at 2,000 g, and the supernatant was collected. Subsequently, sodium chloride (NaCl) was added to a concentration of 0.58 M and incubated at RT for 2 hours to eliminate urinary mucoproteins, including Tamm-Horsfall protein^50,51^. The mixture was then centrifuged again at RT for 30 minutes at 8,000 g, and the supernatant was collected. The sample was filtered using a 0.45 μm filter membrane (Millipore, MA, USA, Catalog# HVLP07625), and then loaded into a concentration device (Amicon® stirred cell, Millipore, MA, USA, Catalog# UFSC40001) and ultrafiltered to a volume of 3–4 ml using a 10 kDa NMW ultrafiltration (UF) disc membrane (Millipore, MA, USA, Catalog# PLGC07610). Next, 200 ml of PBS was added, and the sample was ultrafiltered to a volume of approximately 3–4 ml again, resulting in a concentrated component. The concentrated component was transferred to an ultracentrifuge tube and centrifuged at 150,000 g at 4°C for 150 minutes (SW60Ti, OptimaXE-100, Beckman Coulter, Fullerton, CA). The supernatant was discarded, and the precipitation was resuspended in 350 μl of Dulbecco’s phosphate-buffered saline (DPBS, Beyotime, Catalog# C0221D) containing protease and phosphatase inhibitors (PPICs, Beyotime, Catalog# P1046). This resulted in a uTEV sample.

#### 2.1.2 Isolation of uADEVs

Each uTEV sample was mixed with 50 μl of 3% bovine serum albumin (BSA, Beyotime, Catalog# ST023-50g) and incubated for 1 hour at RT with 4 μl of anti-GLAST (ACSA-1)-biotin antibody (Miltenyi Biotec, Catalog# 130-118-984). Subsequently, 10 μl of streptavidin-agarose resin (Thermo Fisher Scientific, Catalog# 53116) and 40 μl of 3% BSA were added, followed by incubation for 60 minutes at RT. After centrifugation at 800 g for 10 minutes at 4°C and removal of the supernatant, each sample was resuspended in 100 μl of cold 0.1M glycine-HCl (pH = 3.0) by gently mixing for 30 seconds. The suspension was then centrifuged at 4,000 g for 10 minutes at 4°C, and the supernatant was collected. Several drops of 1M Tris-HCl (pH = 8.0, Beyotime, Catalog# ST780-500ml) was added to adjust the pH to 7.0. This resulted in a uGLAST^+^EV sample. For western blotting and protein measurements, mammalian protein extraction reagent (M-PER, Thermo Fisher Scientific, Catalog# 78503) with PPICs was added to each uADEV sample or uTEV sample.

#### 2.1.3 Validation of uADEVs

##### 2.1.3.1 Transmission electron microscopy (TEM)

Similar to our previous ADEV studies^47,52^, the TEM was used to get the image of EVs. Twenty μl of the EV sample was added dropwise to 200-mesh grids and incubated at RT for 10 minutes, then the grids were negatively stained with 2% phosphotungstic acid for 3 minutes, and the remaining liquid was removed by filter paper. Then observed with a HT7800 transmission electron microscope (Hitachi High-Tech Corporation, Tokyo, Japan).

##### 2.1.3.2 Nanoparticle tracking analysis (NTA)

The diameter (nm) and concentration (particles/ml) of EV samples were determined using the ZetaView PMX 110 (Particle Metrix, Meerbusch, Germany) with ZetaView 8.04.02 nanoparticle tracking software (Particle Metrix, Meerbusch, Germany).

##### 2.1.3.3 Western blotting

Western blot was conducted to detect three EV markers with primary rabbit anti_cluster of differentiation (CD)63 antibody (Abcam, Catalog# ab134045), rabbit anti_CD9 antibody (Abcam, Catalog# ab125011), and mouse anti_Alix antibody (Proteintech, Catalog# 67715-1-Ig), an astrocyte marker with rabbit anti-glial fibrillary acidic protein (GFAP) antibody (Abcam, Catalog# ab68428), and two kidney markers Na^+^–K^+^–Cl^−^ cotransporter (NKCC) 2 (Abcam, Catalog# ab171747), sodium-chloride cotransporter (NCC) (Abcam, Catalog# ab95302).

##### 2.1.3.4 Protein measurements

Astrocyte related neurotrophins (brain-derived neurotrophic factor (BDNF), epidermal growth factor (EGF), fibroblast growth factor (FGF)-2, glial cell-derived neurotrophic factor (GDNF), GFAP, nerve growth factor beta (NGF-β), S100 calcium binding protein B (S100B)) were measured using the Human ProcartaPlex™ Simplex kit (Thermo Fisher Scientific, Catalog# PPX-07).

#### 2.1.4 Statistical methods

For comparisons between uTEVs and uADEVs, the concentrations of neurotrophins (pg/ml) were normalized to a reference of 10E+10 particles/ml, yielding values in pg/per 10E+10 particles, adhering to MISEV2018^16^. The fold change of the uADEVs/uTEVs ratios were calculated for both particle and neurotrophin concentrations. Welch’s two sample *t*-tests were employed to test the differences of each parameter between the uADEVs and uTEVs samples. A two-sided *p*-value <0.05 was considered statistically significant. All statistical analyses were performed using R version 4.2.0 (R Project for Statistical Computing) within Rstudio version 1.4.1106 (Rstudio).

### 2.2 Case: the potential ability of uADEVs on depicting the trajectories of target molecules in CNS

To assess the potential of uADEVs in tracking the trajectories of *in vivo* target molecules in the CNS, we analyzed CSF, uADEV, and blood samples from a middle-aged female patient (Patient A) with anti-N-methyl-d-aspartic acid receptor (NMDAR) encephalitis probably caused by teratoma. N-methyl-d-aspartic acid receptor (NMDAR) subunit zeta-1 (GluN1), the pathogenic molecule in anti-NMDAR encephalitis, was measured in CSF and uADEV samples using the enzyme-linked immunosorbent assay (ELISA) (CUSABIO, Catalog# CSB-EL009911HU). Given the patient’s comatose state upon admission, informed consent was obtained from their legal guardian first, and Patient A also provided her own informed consent upon recovery. The wavelet analysis was performed using the *WaveletComp* package^53^ in R version 4.2.0 (R Project for Statistical Computing) within Rstudio version 1.4.1106 (Rstudio).

## 3. Results

### 3.1 Validation of uTEV and uADEVs

Figure 1 (a–h) shows the isolation schematic diagram of uADEVs and their validation using NTA, TEM, and western blotting. NTA confirmed that the EV diameters were within the expected size range for small EVs. TEM images revealed characteristic EV-like structures in both uTEV and uADEV samples. Western blotting showed positive expression of three EV markers (CD63, CD9, and Alix) in both uTEV and uADEV samples. Additionally, uADEVs exhibited positive expression of an astrocyte marker (GFAP). Notably, two kidney markers, NCC2 and NKCC, were detected in the uTEV sample but not in the uADEVs samples. See **Supplementary Material 1 (s**Figure 1) for the original western blotting images.

### 3.2 Comparisons between uTEVs and uADEVs

Particle concentrations in uADEVs (5.3 ± 1.6E10/ml) were significantly lower compared to uTEVs (1.9 ± 0.78E12/ml). Given the sample volumes of uADEVs (216 µl) and uTEVs (900 µl), this indicates that uADEVs constitute about 0.81% of uTEVs. In contrast, neurotrophin levels in uADEVs were notably higher compared to uTEVs. To quantify the enhancement in astrocytic signal clarity within the CNS, we assessed the signal-to-noise ratio (SNR) by calculating the fold increase in neurotrophin concentrations. This analysis revealed a range of fold changes between 23.1 and 88.1 across seven neurotrophins, as detailed in Figure 1(i) and **Table 1**.

**Table 1.** Fold changes of uADEVs to uTEVs.

| Variable | In uADEV (pg)<sup>a</sup> | In uTEV (pg)<sup>a</sup> | Increased Fold Change |  | *p* |
| --- | --- | --- | --- | --- | --- |
|  | Mean ± SD | Mean ± SD | Mean ± SD | Median [IQR] |  |
| BDNF | 0.625 ± 0.246 | 0.010 ± 0.005 | 88.1 ± 69.3 | 61.0 [33.8; 112.1] | <0.001 |
| EGF | 38.153 ± 9.364 | 2.761 ± 2.126 | 23.1 ± 18.1 | 17.6 [8.3; 32.6] | <0.001 |
| FGF-2 | 3.747 ± 1.991 | 0.162 ± 0.124 | 28.8 ± 16.5 | 29.9 [19.9; 30.6] | 0.001 |
| GDNF | 23.355 ± 4.966 | 0.897 ± 0.685 | 32.8 ± 11.4 | 31.0 [28.8; 44.0] | <0.001 |
| GFAP | 7.826 ± 6.890 | 0.285 ± 0.203 | 29.9 ± 17.3 | 29.0 [15.0; 33.6] | 0.011 |
| NGF-β | 0.985 ± 0.879 | 0.041 ± 0.028 | 40.4 ± 47.5 | 13.0 [9.6; 80.7] | 0.012 |
| S100β | 0.765 ± 0.219 | 0.029 ± 0.021 | 43.7 ± 34.6 | 35.4 [17.8; 59.9] | <0.001 |

Note: a: Normalized to every 10<sup>10</sup> particles.

### 3.2 Case: the longitudinal trajectory of proteins in uADEVs in an encephalitis patient

Figure 2 demonstrated the comprehensive dynamic picture of Patient A during hospitalization.

**Figure 2.**
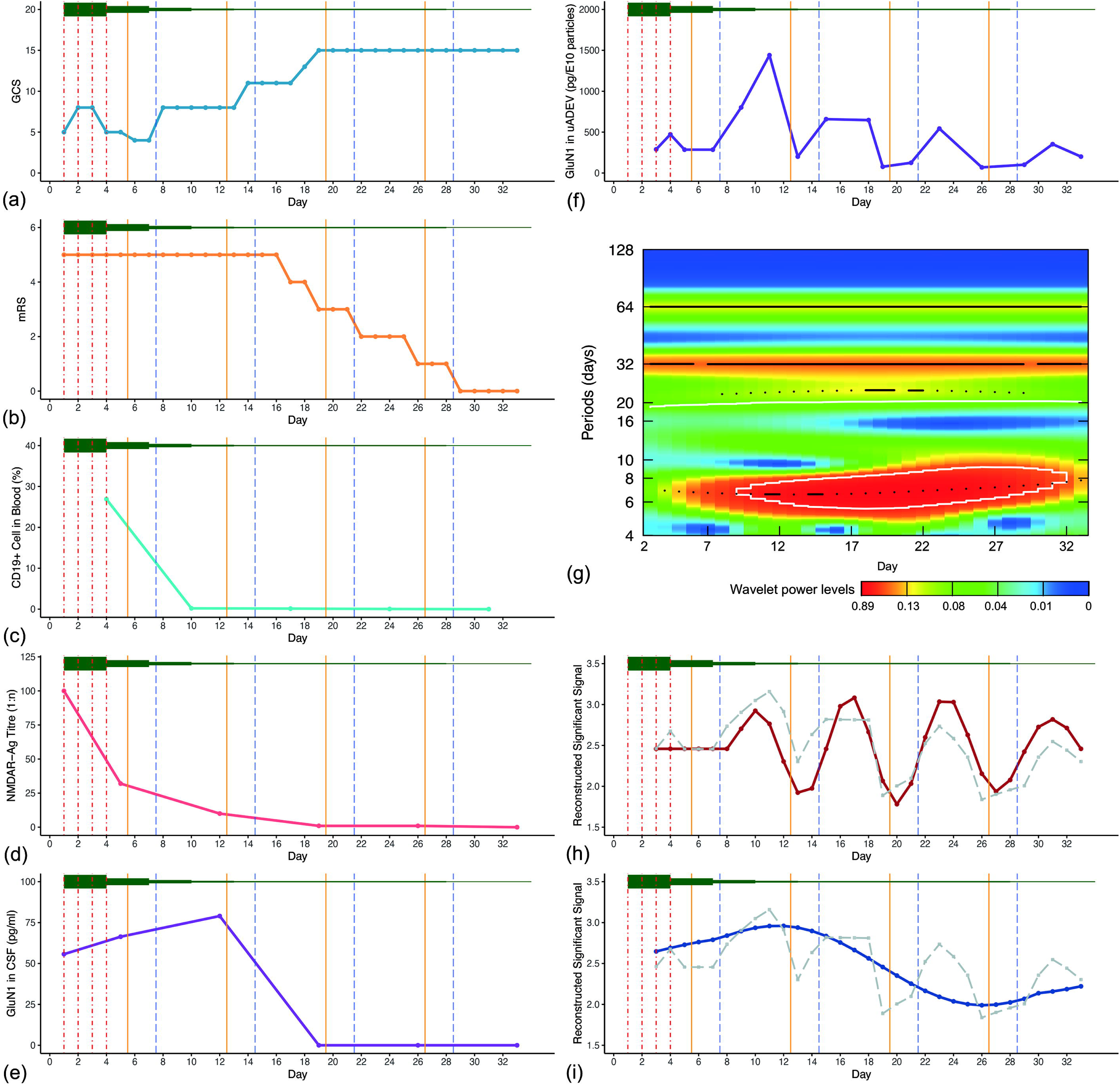
Comprehensive dynamic picture of Patient A with anti-NMDAR encephalitis during her hospitalization. The x-axis in all panels marks the days since admission (Day 0 = admission day). Main treatment included an initial phase of four intravenous infusions of human γ-globulin (dose 25 g, indicated by 4 vertical dot-dash red lines). Simultaneously, glucocorticoid therapy was administered (represented by the green horizontal line above each subfigure, with the line’s width indicating the equivalent dose of methylprednisolone). Additionally, intrathecal methotrexate administration (dose 10 mg, represented by vertical solid yellow lines) and rituximab therapy (dose 100 mg, indicated by vertical long-dash blue lines) were administered. Clinical symptoms were assessed using two scales: (a) the Glasgow Coma Scale (GCS) and (b) the modified Rankin Scale (mRS), showing gradual improvement following treatment. Given the use of the immunotherapy, we measured the (c) percentage of CD19+ cells in the blood, assessed via flow cytometry, dropped to nearly non-existent levels by Day 10. We also monitored NMDAR-antigen (Ag) titres in the CSF. (d) A decrease in NMDAR-Ag titers in the CSF was observed as the treatment progressed. Given that anti-NMDA encephalitis is characterized by the presence of autoantibodies mainly against the NMDAR GluN1 subunit, leading to NMDARs damage through internalization, shedding, and extracellular release, we also measured (e) The concentrations of GluN1 in CSF. As NMDARs are also present on the surface of astrocytes, we simultaneously measured the (f) concentrations of GluN1 in uADEVs (pg/per 1E+10 particles). As the trajectory of GluN1 in uADEVs was also a dynamic signal, composed of various wave components. Therefore, we conducted wavelet analysis using the Morlet wavelet as mother wavelet to identify and separate the significant components of this dynamic signal. (g) Wavelet analysis results of the log(10) GluN1 trajectory in uADEVs are presented, with colors representing power, black lines indicating the highest peak (ridge) in the respective region, and white areas representing significant components. Two significant components were identified, one with a short period (approximately 6-8 days) located below and another with a period of over 20 (the strongest ridge is at approximately 32 days). These two significant components were then reconstructed as follows: (h) The trajectory of the reconstructed significant short-period component (red line), and (i) the long-period component (approximately 32 days, the blue line). The gray dashed line in (h) and (i) represent the original log(10) trajectory (grey dashed lines).

### 3.3 Data availability

All data were presented in the **Supplementary Material 2: Individual Participant Data**.

## 4. Discussion

In this study, we established a method to extract ADEVs from urine, facilitating the tracing of *in vivo* specific molecular signal in the CNS. Then, as a demonstration, we tracked the trajectory of NMDAR subunit GluN1 in uADEVs from a patient with anti-NMDAR encephalitis and employed wavelet analysis to identify significant components in the trajectory. This uADEVs protocol may offer a novel, non-invasive method for daily CNS monitoring, providing a valuable tool for biomarker discovery and etiological studies of rapidly evolving CNS diseases.

In order to verify the isolation efficiency of uADEVs, we compared the neurotrophin concentrations normalized to particle numbers in uTEV and uADEV samples. Mean fold changes ranged from 23.1 (EGF) to 88.1 (BDNF) (Figure 1(i)). This enrichment can be interpreted from the signal-theoretic perspective as an improvement in the SNR of astrocyte-derived signals in uTEVs.

To illustrate the concept of signal amplification, we consider a scenario where “a” represents the total number of EVs extracted from a urine sample, and “b” represents the number of EVs are derived from astrocytes (uADEVs). The original SNR of the signal from astrocytes in uTEVs is 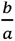, and the SNR of the same signal in uADEVs is 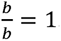. Therefore, the amplification factor is 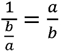, which is approximately 123 based on particle number ratio in this present study. This indicates that the SNR of a signal from astrocytes in uGLAST^+^EVs is 123 times higher than in uTEVs, representing an upper bound for the increase in SNR. The increase in SNR was also assessed using astrocyte-related molecules, such as neurotrophins. The fold increase in the ratio of their concentration to particle number in uADEV samples compared to uTEV samples provides an estimate of the lower bound of SNR enhancement. The results revealed a maximum 88-fold increase (BDNF), which is slightly lower than the estimate based on particle count. Our approach, therefore, leads to an approximately 88–123-fold increase in SNR for signals from astrocytes via uADEVs. Nevertheless, these are preliminary estimates, and further studies are warranted to refine these calculations.

Another key issue of this protocol is to estimate potential contamination mainly from urogenital tract. Hence, we selected two widely used markers of uTEVs (NKCC2 and NCC) in urinary EV studies^31,54^. Western blotting results showed that these two markers were highly expressed in uTEV samples as expected, but were barely detected in uADEV samples (Figure 1(h)). These results indicate minimal kidney-derived contamination in uADEVs. Along with evidence of neurotrophins, we established that the majority of these uGLAST^+^EVs were ADEVs.

Theoretically, uTEVs may also contain EVs derived from other CNS cells, such as neurons and various types of glial cells, including microglia and oligodendrocytes, which could be isolated using similar methods. For other types of BDEVs, such as NDEVs, L1 cell adhesion molecule (L1CAM) is commonly used as a marker^55–57^. However, L1CAM expression is also detected in the kidney^58^, potentially compromising the purity of urinary L1CAM^+^EVs that are truly derived from CNS. Given that GLAST is believed to be predominantly expressed in astrocytes, we selected urinary GLAST^+^EV/ADEV as an initial proof-of-concept for isolating BDEVs from urine. We are developing appropriate methodologies to exclude the interference of EVs from the urological system, enabling the enrichment of other types of BDEVs from urine.

UC has been extensively used for uTEV enrichment for decades, with a common protocol involving UC at 100,000 g for 70 minutes, repeated twice, to isolate uTEVs ^59–65^. Investigations indicated that extended centrifugation, potentially up to four hours, may enhance EV yields^66^. In our study, given the low proportion of uADEVs in uTEVs, our primary objective was to increase the yield of uTEVs rather than focusing on purity at the first stage. Therefore, we conducted a pilot study to refine UC parameters for optimal uTEVs yield, detailed in **Supplementary Material 3 (The Determination of the Duration of Ultracentrifugation)**, and the TEM images (Figure 1**(c and d)**) suggested that the applied UC parameters (150,000 g for 150 minutes) did not significantly introduce impurities.

We also presented longitudinal data from a patient with anti-NMDAR encephalitis to showcase the potential of uADEVs in revealing *in vivo* molecular trajectories and demonstrate the application of signal processing algorithms for enhanced analysis (Figure 2). Anti-NMDAR encephalitis is a well-defined autoimmune disease characterized by autoantibodies targeting the NMDA receptor (primarily the GluN1 subunit) in the CNS^67,68^. The first-line immunosuppressive therapy typically comprising steroids, intravenous immunoglobulin, and plasma exchange, followed by second-line therapies including rituximab or methotrexate/cyclophosphamide, alone or in combination^69,70^. In this longitudinal data, we observed dynamic changes in GluN1 within uADEVs. To analyze these dynamic changes, we applied wavelet analysis, a widely used algorithm that decomposes complex signals into simpler components^71^. This approach enables us to identify and characterize the underlying patterns in GluN1 fluctuations. We discovered that it consists of two main significant components, with the shorter periodic component (approximately 6–8 days) shown in Figure 2(h) and the longer one (approximately 32 days) in Figure 2(i). Based on prior knowledge, here we made some guesses. Since some EVs are formed through cell membrane invagination, and inevitably carry extracellular molecules^72–74^. Additionally, the shape and time course of this component resemble the trajectory of GluN1 in CSF (Figure 2(e)), we speculate that the longer-period component may reflect the changes in extracellular GluN1 levels. The shape and timing of the shorter period component appeared to correlate with the treatment. Following the combination of methotrexate and rituximab, the trajectory of the short-period component increased. Considering the plasma membrane origin of EV membranes and previous reports of NMDAR density reduction in anti-NMDAR encephalitis patients and its restoration upon effective treatment^75^, we hypothesize that this short-period signal may reflected the dynamic response of NMDAR density on astrocyte plasma membranes to treatment. However, it is important to note that definitive conclusions cannot be drawn from a single case in this study, the aforementioned conjectures are more suitable as new hypotheses for further investigation. Our study suggests that by increasing the number and density of sampling points, sophisticated powerful signal processing algorithms like wavelet analysis could be applied to extract meaningful information from these dynamic CNS *in vivo* signals, enabling the formulation of more specific hypotheses for targeted experimental validation.

## Significance

The temporal depth provided by uADEVs may hold significant promise for the discovery of novel biomarkers for CNS diseases, particularly those that progress rapidly. One prerequisite for a molecule (or a combination of molecules) to become a reliable diagnostic marker, is that it must be “individual-level” identifiable. Nevertheless, for rapidly evolving CNS diseases, conventional follow-up frequencies (monthly, semiannually, or annually) may not be sufficient to capture the dynamic molecular changes associated with disease progression, according to the Nyquist-Shannon sampling theorem^15^ (Figure 3(a)). However, our uADEVs protocol could offer a non-invasive solution with the potential for sampling up to daily. With advancements in detection technology and reduced urine volume requirements, hourly sampling may become feasible, enabling the capture of CNS dynamics at an unprecedented level. In short, uADEVs allow us to capture *in vivo* molecular “movies” of the CNS at the individual level, rather than capturing static “snapshots”. This further implies that, diagnosing a disease with distinct episodes may require multiple longitudinal tests, even including artificial perturbations (analogous to an oral glucose tolerance test). This is because a single-time biomarker test may not be sufficient to capture the dynamic nature of such diseases.

**Figure 3.**
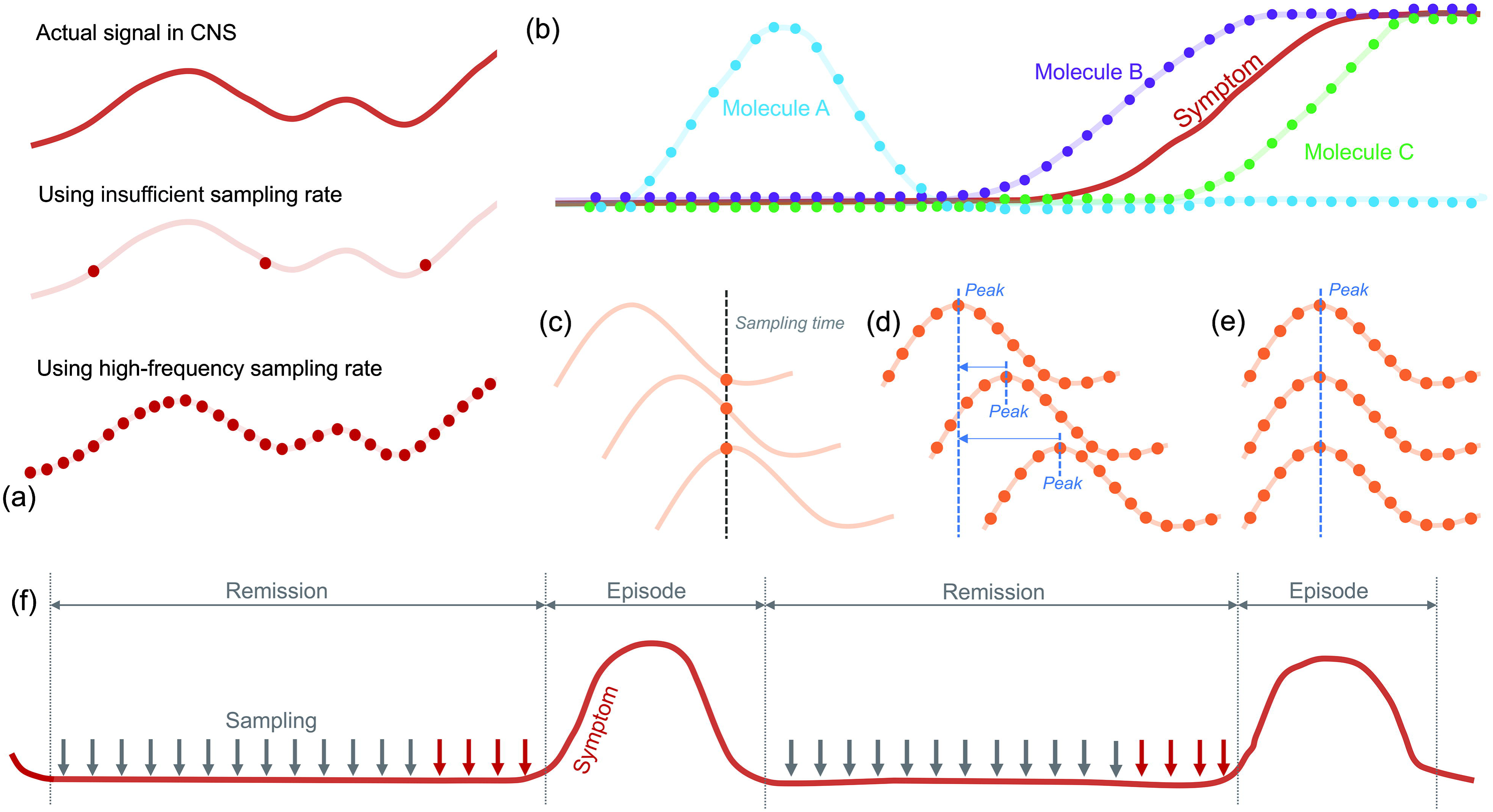
The Significance of the uADEV approach. (a) The advantage of high-frequent sampling. (b) The detailed trajectories of target molecules may benefit for the exploration (Molecule A and B) and falsification (Molecule C) of pathological hypotheses. (c–e) Even under the assumption that all patients have the same molecular trajectories, yet, time-induced heterogeneity exist merely due to different sampling points along the trajectory. However, individual-level detailed trajectories may allow some *post-hoc* algorithms, such as peak-based realignment, to reduce the time-induced heterogeneity. (f) The schematic diagram of the “Time Machine of Sampling”: for CNS diseases with a high probability of recurrence, we could collect urine samples at a high frequency and store them. When a patient is monitored for a recurrence, we could then unseal the samples before the recurrence time point (red arrows) to extract uADEVs to explore the reason of recurrence, like the “Time Machine”.

Even if no new biomarkers are discovered, uADEVs will facilitate the falsification of potential hypotheses. As illustrated in Figure 3(b), if the trajectory of a candidate molecule aligns with, but lags behind, the symptom trajectory, it can be inferred that it is not the cause but rather a consequence or a confounding factor. Conversely, only candidate molecules with trajectories preceding the symptom trajectory are likely to be causal. Naturally, things become somewhat simpler when we can precisely trace the molecular trajectories at the individual-level.

Identifying reliable biomarkers for rapidly changing CNS disorders, such as depression, is challenging, potentially due to their high heterogeneity, and the fluctuating molecular signals themselves might also contribute to this heterogeneity. As illustrated in Figure 3(c), even assuming all patients with a specific disease share identical molecular trajectories (this assumption is inaccurate but serves to demonstrate the concept), heterogeneity can also arise from different sampling points along the trajectory. This time-induced heterogeneity could be a significant factor. However, with adequate sampling frequency, we can capture individual molecular trajectories (Figure 3(d)). Various *post-hoc* algorithms, such as realigning trajectories based on their peaks, can then be employed to reduce time-induced heterogeneity (Figure 3(e)). While this is a simplified model, and heterogeneity manifests in various forms with greater complexity in real-world data, we believe that high-frequency sampling provides more opportunities for data processing using multiple algorithms, enabling deeper exploration.

This methodology may introduce a novel experimental paradigm, which we termed the “Time Machine of Sampling” (Figure 3(f)). Studying the recurrence of CNS diseases is often hampered by the limited availability of samples from the period preceding recurrence. These samples are crucial as they may contain the molecular trigger signals for recurrence. Fortunately, uADEVs can be non-invasively collected and directly reflect CNS signals. For CNS diseases with a high recurrence probability, we propose collecting and storing urine samples at a high frequency. When monitoring a patient for recurrence, we can unseal samples collected before the recurrence time point to extract uADEVs for analysis. This may enable the collection of samples with a sufficient sampling rate from the period preceding CNS disease recurrence. Hence, we term this paradigm the “Time Machine of Sampling” as it allows us to go back in time and collect samples before recurrence. This paradigm may facilitate the identification of molecular triggers preceding disease recurrence.

Our study may also enhance therapeutic monitoring. Timely assessment of treatment efficacy is essential for precision medicine. As demonstrated in Patient A, the GluN1 concentrations in uADEVs appear to be responsive to treatment. This suggests that the uADEVs approach could provide more rapid and timely feedback for diseases that demand close therapeutic monitoring. Further research in this area is warranted.

It is noteworthy that, based on the protocol’s principle, it is theoretically possible to isolate not only ADEVs from the CNS but also EVs from other CNS cell types, organs, or tissues from uTEVs, if these cells express specific surface markers. We anticipate that urine samples may hold significant value for disease studies beyond CNS and urological disorders.

## Limitations

First, alternative isolation protocols, such as size-exclusion chromatography (SEC), may be suitable for laboratories lacking ultracentrifuges. Additionally, UF may lead to a significant loss of uTEVs. Employing high-capacity ultrafiltration tubes to directly collect uTEVs without UF may potentially enhance the yield of uADEVs. However, due to laboratory constraints, we were unable to attempt this, and further investigation is required by external laboratories. Second, while the yield of uTEVs is substantial, the absolute number of uADEVs remains low, limiting multi-omics-based high-throughput assays. To address this challenge, we are endeavoring to develop methodologies for high-throughput studies utilizing minimal amounts of uADEVs. Third, the impact of subjects’ disease status, particularly urological diseases, on uADEVs remains unclear. Further research is needed to address this question. Fourth, the mechanism by which ADEVs traverse the glomerular basement membrane into urine remains unknown. Elucidating this mechanism may significantly enhance the utility of uADEVs. Fifth, this study utilized fresh urine samples. The applicability of frozen or concentrated urine samples after thawing remains undetermined. Future studies should investigate the effect of sample storage conditions on uADEVs. After all, compared with unconcentrated urine samples, storing concentrated urine samples could alleviate inventory pressure on biobanks.

## Conclusions

In this study, we proposed a simple method for isolating urinary ADEVs, enabling non-invasive monitoring of CNS *in vivo* activity with high sampling rates, up to daily or even half-daily. This approach, analogous to capturing molecular “movies” of the brain, coupled with appropriate signal processing algorithms, holds promise for identifying novel biomarkers or illuminating the etiology of rapidly evolving CNS diseases by tracking the precise trajectories of target molecules.

## Supporting information

Supplementary Material 1: sFigure 1. The original whole piece pictures of western blotting.

Supplementary Material 2: Individual Participant Data.

Supplementary Material 3: The Determination of the Duration of Ultracentrifugation.

## Data Availability

All data were presented in the Supplementary Material.

## Author Contributions

Xin-hui Xie: Conceptualization, Methodology, Formal analysis, Investigation, Writing - Original Draft, Writing - Review & Editing, Visualization, Supervision. Mian-mian Chen: Investigation, Methodology, Writing - Original Draft. Shu-xian Xu: Methodology, Investigation, Writing - Original Draft. Jun-hua Mei: Investigation, Writing - Original Draft. Qing Yang: Investigation, Writing - Original Draft. Chao Wang: Methodology, Writing - Review & Editing. Zhongchun Liu: Funding Acquisition, Project Administration, Supervision.

## Acknowledgement

This work was supported by grant from the National Natural Science Foundation of China (grant number: U21A20364). This work has not received funding/assistance from any commercial organizations. The funding source had no roles in the design of this study and will not have any roles during the execution, analyses, interpretation of the data, or decision to submit results.

## Supplementary Materials

**Supplementary Material 1: sFigure 1. The original whole piece pictures of western blotting**. GFAP: Anti-GFAP antibody (Abcam, Cat No: ab68428) at 1/5000 dilution, protein loading amount: 13.7 μg Per lane. NKCC2: Anti-NKCC2 antibody (Abcam, Cat No: ab171747) at 1/2000 dilution, protein loading amount: 4 μg per lane. NCC: Anti-NCC antibody (Abcam, Cat No: ab95302) at 1/1000 dilution, protein loading amount: 1.5 μg per lane. CD63: Anti-CD63 antibody (Abcam, Cat No: ab134045) at 1/700 dilution, protein loading amount: 4.0 μg per lane. CD9: Anti-CD9 antibody (Abcam, Cat No: ab263019) at 1/700 dilution, protein loading amount: 5.7 μg per lane. Alix: Anti-Alix antibody (proteintech, Cat No: 67715-1-Ig) at 1/2000 dilution, protein loading amount: 5.7 μg per lane.

**Supplementary Material 2: Individual Participant Data.**

**Supplementary Material 3: The Determination of the Duration of Ultracentrifugation.**

