## Supplementary figures and images for "Urinary Astrocyte-derived Extracellular Vesicles: A Non-invasive Tool for Capturing Human *In Vivo* Molecular “Movies” of Brain"

### Supplementary Material 1: sFigure 1. The original whole piece pictures of western blotting.

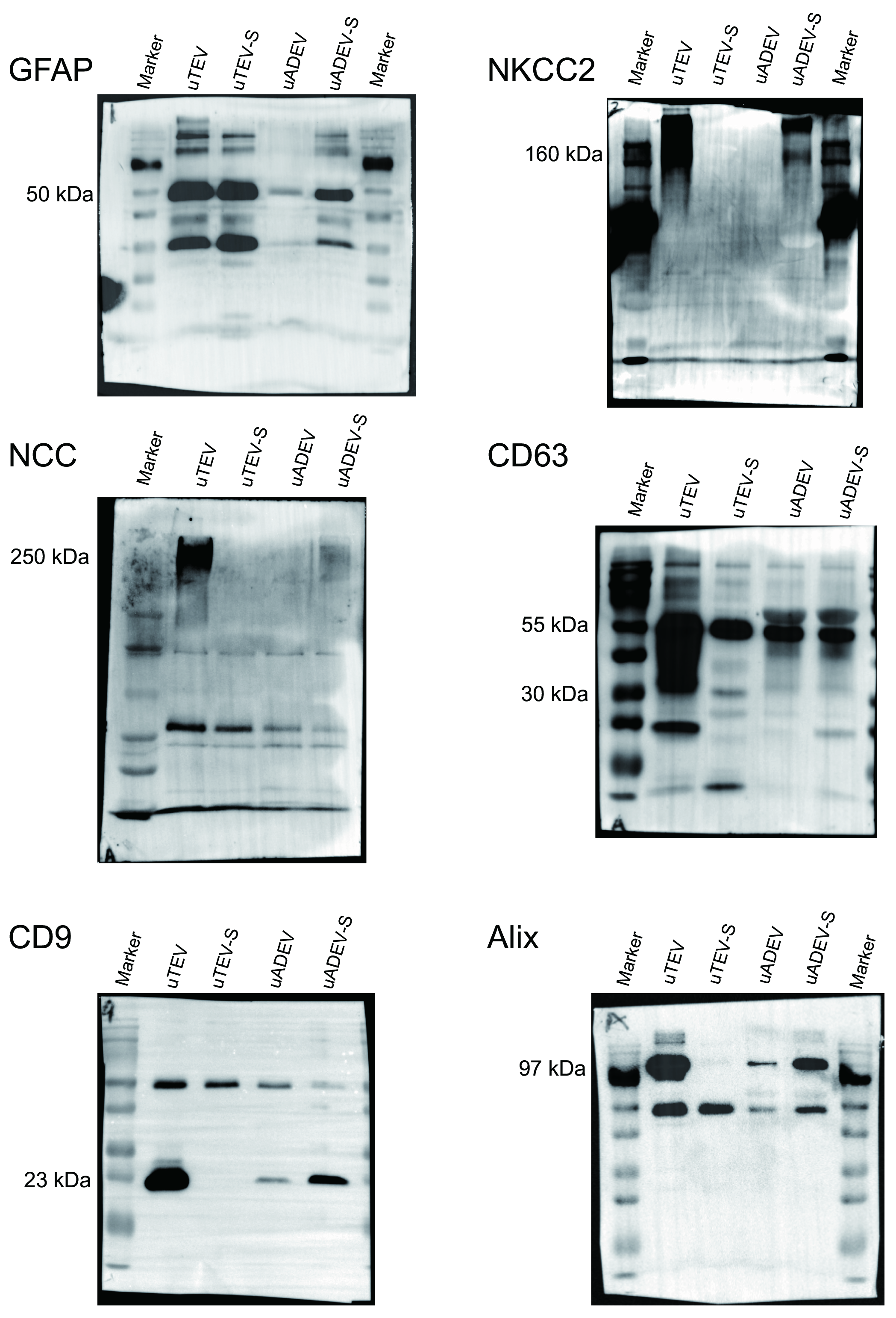
