## Supplementary Material 3: The Determination of the Duration of Ultracentrifugation. for "Urinary Astrocyte-derived Extracellular Vesicles: A Non-invasive Tool for Capturing Human *In Vivo* Molecular “Movies” of Brain"

**Supplemental material: The Determination of the Duration of Ultracentrifugation.**

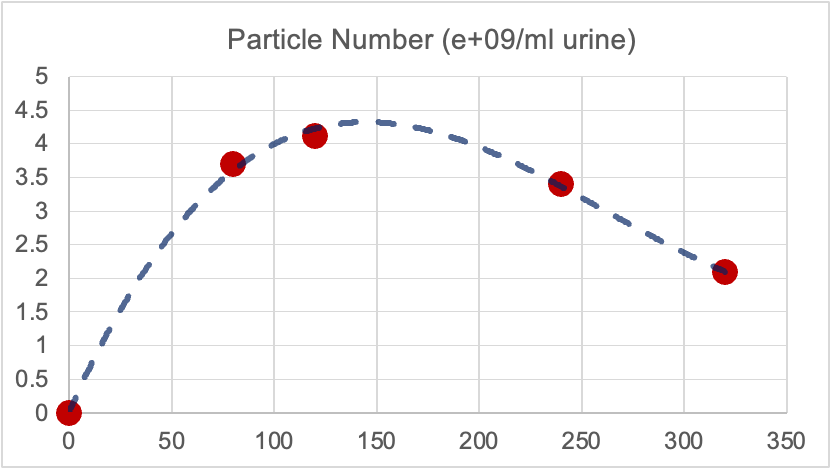
In this study, it is important to maximize the yield of urinary total extracellular vesicles (uTEVs). Therefore, we conducted the following tests to investigate the impact of varying centrifugation times on the yield of uTEVs from concentrated urine samples, while employing the same preprocessing steps and centrifugation force (150,000 g). The results are presented in the table and figure below:

| Time (min) | Particle number/ml urine |
| --- | --- |
| 80 | 3.70E+09 |
| 120 | 4.12E+09 |
| 240 | 3.4E+09 |
| 320 | 2.09E+09 |

Therefore, we determined the ultracentrifugation time is 150 minutes at 150,000 g.
